# Autoantibody production is enhanced after mild SARS-CoV-2 infection despite vaccination in individuals with and without long COVID

**DOI:** 10.1101/2023.04.07.23288243

**Authors:** L Visvabharathy, C Zhu, ZS Orban, K Yarnoff, N Palacio, M Jimenez, PH Lim, P Penaloza-MacMaster, IJ Koralnik

## Abstract

Long COVID patients who experienced severe acute SARS-CoV-2 infection can present with humoral autoimmunity. However, whether mild SARS-CoV-2 infection increases autoantibody responses and whether vaccination can decrease autoimmunity in long COVID patients is unknown. Here, we demonstrate that mild SARS-CoV-2 infection increases autoantibodies associated with systemic lupus erythematosus (SLE) and inflammatory myopathies in long COVID patients with persistent neurologic symptoms to a greater extent than COVID convalescent controls at 8 months post-infection. Furthermore, high titers of SLE-associated autoantibodies in long COVID patients are associated with impaired cognitive performance and greater symptom severity, and subsequent vaccination/booster does not decrease autoantibody titers. In summary, we found that mild SARS-CoV-2 infection can induce persistent humoral autoimmunity in both long COVID patients and healthy COVID convalescents, suggesting that a reappraisal of vaccination and mitigation strategies is warranted.

## Introduction

The long-term clinical impact of SARS-CoV-2 infection remains unclear. More than 100 million people in the United States have contracted SARS-CoV-2 infection, and of these approximately one third of survivors report post-infectious sequelae of SARS-CoV-2 infection (PASC), or long COVID, at 4 months^1, 2^. PASC patients frequently complain of fatigue, myalgia, brain fog, and other neurologic sequelae as the primary drivers of decreased quality of life^3^. Although mRNA vaccines against SARS-CoV-2 have been extremely effective in preventing severe acute disease^4^, incidence of PASC has not significantly decreased in the US despite widespread vaccine uptake^5^. This indicates that PASC will remain a medical concern for the foreseeable future.

Many comprehensive studies have found associations between humoral autoimmunity and severe acute COVID-19 and PASC. Patients hospitalized with severe or moderate COVID-19 were significantly more likely to exhibit autoantibody responses against cytokines, chemokines, and immune co-receptors^6^ or G protein-coupled receptors^7^ compared to those with mild or asymptomatic acute presentation. Similarly, the development of anti-nuclear autoantibodies 2-3 months after acute SARS-CoV-2 infection in hospitalized patients^8^ and in those with moderate to severe acute symptoms^9^ had predictive value in identifying PASC patients while associating with persistent fatigue, cough, and dyspnea. Importantly, high intrathecal and serum levels of anti-dsDNA autoantibodies were found in patients with severe neurologic symptoms including dizziness, neuropathy, myopathies, and encephalopathy during acute infection^10^. The common thread between these studies lies in the association of elevated autoantibody responses with severe acute disease. However, we and others have also demonstrated that PASC occurs just as frequently in patients with mild acute disease^11–13^, and it has been suggested that SARS-CoV-2 vaccines can decrease the severity of long COVID^14^, albeit moderately^5^. This brings up two critical questions: does humoral autoimmunity contribute to PASC symptoms in patients who experienced mild acute COVID-19, and does vaccination decrease autoantibody titers after infection in PASC patients?

Here, we longitudinally track vaccine-elicited and autoantibody responses in PASC patients with primarily neurologic symptoms (Neuro-PASC) and COVID convalescent controls. Our data show three critical findings linking SARS-CoV-2 infection, autoantibody responses, and Neuro-PASC. First, mild acute SARS-CoV-2 infection results in vigorous increases in autoantibody titers in Neuro-PASC patients and to a lesser degree in COVID convalescents without persistent symptoms. Second, antibodies linked to systemic lupus erythematosus (SLE) and inflammatory myopathies are enriched in Neuro-PASC patients and titers are not decreased after receiving a booster dose of mRNA vaccines. Third, elevations in anti-cytokine, neuronal, and anti-nuclear autoantibodies in Neuro-PASC patients are strongly associated with cognitive dysfunction and increased severity of neurologic symptoms. Together, these data indicate that mild SARS-CoV-2 infection results in sustained elevation of autoantibodies in both Neuro-PASC patients and to a lesser extent in those without persistent symptoms, highlighting the need for reappraisal of mitigation strategies to prevent infection.

## Results

### Clinical characteristics of study participants

We enrolled a total of 46 study participants, all of whom had been vaccinated with 2 doses of the primary series of mRNA-1273 (Moderna) or BNT162B2 (Pfizer) SARS-CoV-2 mRNA vaccines prior to enrollment. Participants were recruited from the Neuro-COVID-19 outpatient clinic at Northwestern Memorial Hospital or from the surrounding Chicago area. These included 17 Neuro-PASC patients (“NP”; confirmed RT-PCR^+^ or rapid antigen^+^ before or after vaccination, or seropositive prior to vaccination) recruited between August 2021-March 2022 and who had neurologic symptoms lasting at least 8 weeks but up to 88 weeks post-infection. Among those, 16 (94.1%) had mild acute disease without pneumonia or hypoxia and not requiring hospitalization. We additionally recruited 14 COVID convalescent controls (“CC”; RT-PCR^+^ or rapid antigen^+^ pre- or post-vaccination) without symptoms persisting for more than 4 weeks from onset, all of whom had mild acute disease. This group was intentionally selected to be younger and with fewer comorbidities than the NP group to limit the effects of older age and comorbidities on autoantibody titers pre-infection. Finally, 15 healthy controls with no prior history of SARS-CoV-2 exposure or autoimmune disease (“HC”, RT-PCR^-^ and seronegative) were also included. Plasma samples were collected pre-boost, 3 weeks post-boost, and 3.5 months post-boost to assess longitudinal vaccine elicited and autoantibody responses (study design in Fig. 1A).

**Figure 1:**
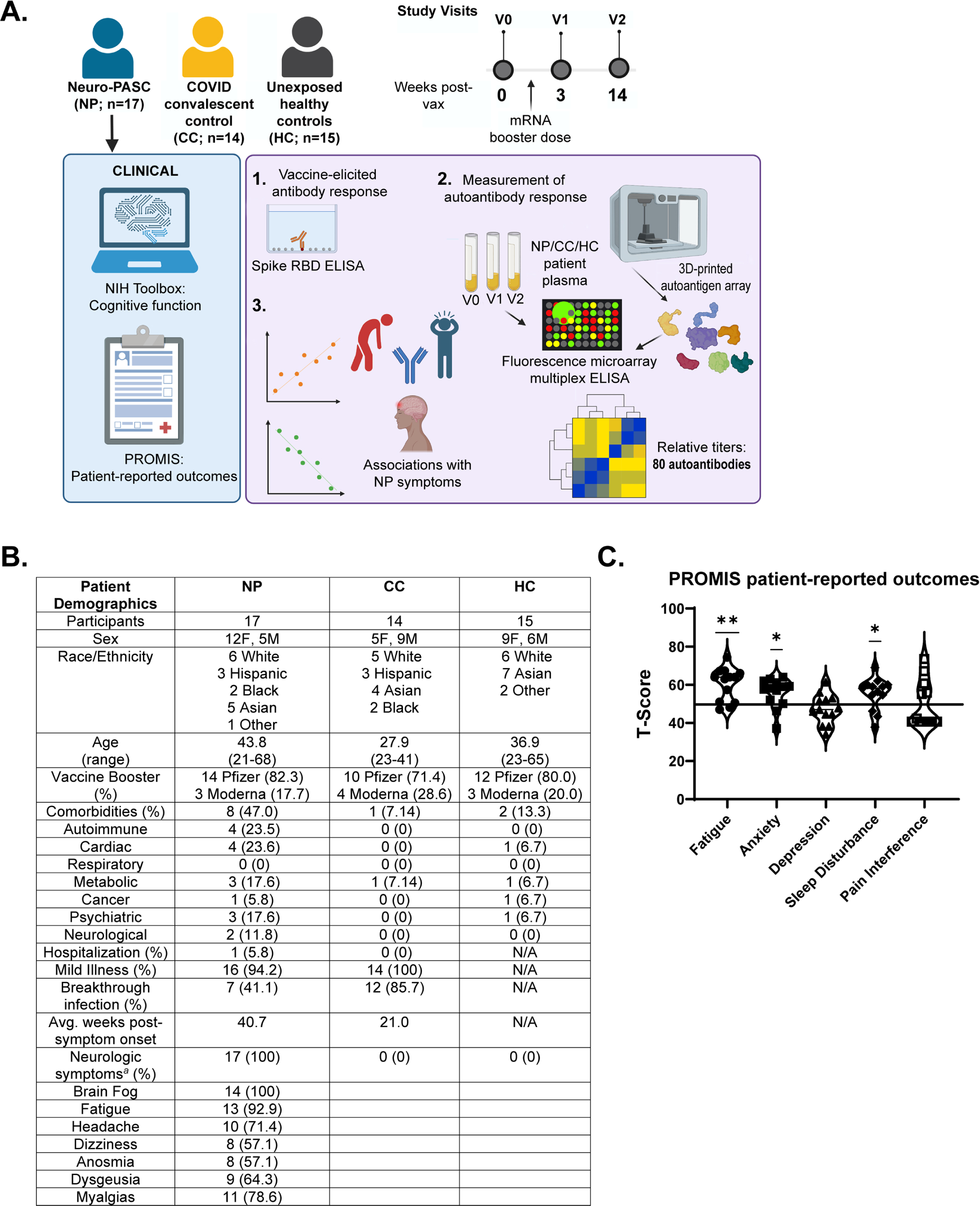
Study design and patient demographics. A.) Longitudinal study design. B.) Demographic table for Neuro-PASC (NP), convalescent controls (CC), and healthy control (HC) study participants. C.) PROMIS patient-reported outcome survey data for NP patients. Horizontal black line represents the U.S. national average T score of 50; *p* values relative to demographic-matched US national average by one sample t test. *p<0.05; **p<0.01. *^a^* indicates the 14/17 NP patients for whom detailed symptom information was collected in the clinic.

Neuro-PASC patients exhibited symptoms consistent with those previously reported by our group^15^, including brain fog, fatigue, headache, and myalgia. Plasma samples were collected at an average of 21-40 weeks post-infection, with 41% of Neuro-PASC and 85.7% of convalescent controls having contracted SARS-CoV-2 as a breakthrough infection post-vaccination (Fig. 1B). Neuro-PASC patients reported quality of life impairment in multiple domains including fatigue, anxiety, and sleep disturbance which were significantly higher than the demographic-matched US national average (Fig. 1C) as shown in other studies^16^.

### Impacts of mRNA vaccination on antiviral immunity and inflammation

Endpoint titer ELISA was used to measure vaccine-elicited antibody responses over time. Spike receptor binding domain (RBD) responses were significantly higher pre-boost in Neuro-PASC and convalescent control subjects with prior SARS-CoV-2 exposure, but all groups exhibited similar titers at 3 weeks and 3.5 months post-boost (Fig. 2A). We next determined whether vaccination increased acute clinical markers of inflammation as previously reported for systemic cytokine levels^17^ by measuring levels of C reactive protein (CRP). There were no significant differences in plasma concentrations of CRP between groups across all timepoints (Fig. 2B).

**Figure 2:**
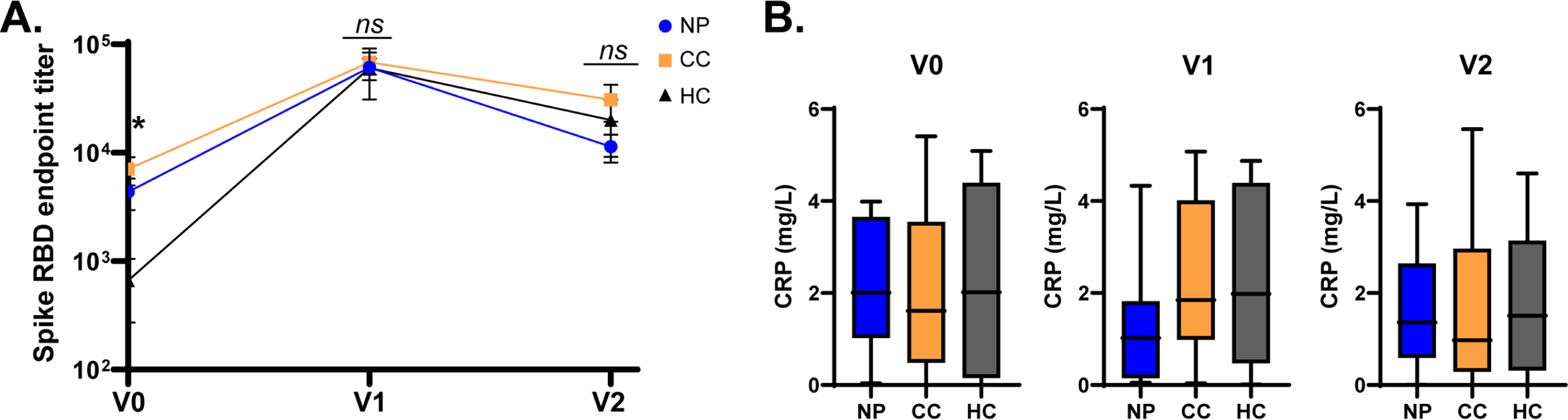
Vaccine-elicited Spike RBD responses are unaffected by long COVID, but NP patients have elevated plasma markers of inflammation post-vaccination. A.) mRNA vaccines elicit similar levels of anti-Spike RBD antibodies in NP, CC, and HC subjects. B.) CRP was not significantly elevated (>10mg/L) in any group across all study visits. *p<0.05; **p<0.01 by one-way ANOVA (A).

### Autoantibodies of diverse specificity are elevated in people with prior SARS-CoV-2 exposure

We next probed autoantibody responses in patients with or without prior SARS-CoV-2 infection before and after their mRNA vaccine booster dose. Plasma samples were collected at the indicated timepoints and screened for the presence of 80 autoantibodies using the IgG Autoantigen Microarray Super Panel from the Immune Monitoring Core at the University of Texas Southwestern Medical Center. Of 17 Neuro-PASC patients, 15 (88%) exhibited polyautoantibody responses (significant elevations in >2 autoantibodies) related to multiple autoimmune diseases or syndromes such as rheumatoid arthritis, systemic sclerosis, and autoimmune vasculitis at V2. Surprisingly, 12 of 14 (85%) of convalescent controls also showed significant elevation in more than 2 autoantibodies with different specificities at all timepoints compared with healthy controls (Fig. 3A, S1). Autoantibodies implicated in inflammatory myopathies and SLE were enriched among those elevated in Neuro-PASC patients and to a lesser extent in convalescent controls when compared to healthy control subjects (Fig. 3B). In addition, anti-cytokine and the neuronal antigen specific α-GAD65 antibodies were also more elevated in Neuro-PASC patients (Fig. S2). The highest autoantibody titers in patients with prior SARS-CoV-2 infection were found at either at V1 (3 weeks post-vaccination) or at V2 (3.5 months post-vaccination) rather than at the baseline (Fig. 3C), though differences were not statistically significant. Autoantibody titers were not significantly affected by age, sex, or time elapsed from acute infection in either group, except for MDA5 autoantibody (Fig. S3). Crucially, acute-phase measures of inflammation such as CRP or acute phase COVID-19 disease severity did not predict whether a study subject would exhibit elevated poly-autoantibody responses (Fig. S4). Overall, these results suggest that even mild prior infection with SARS-CoV-2 can enhance humoral autoreactivity to a greater extent in long COVID patients than in those without persistent symptoms.

**Figure 3:**
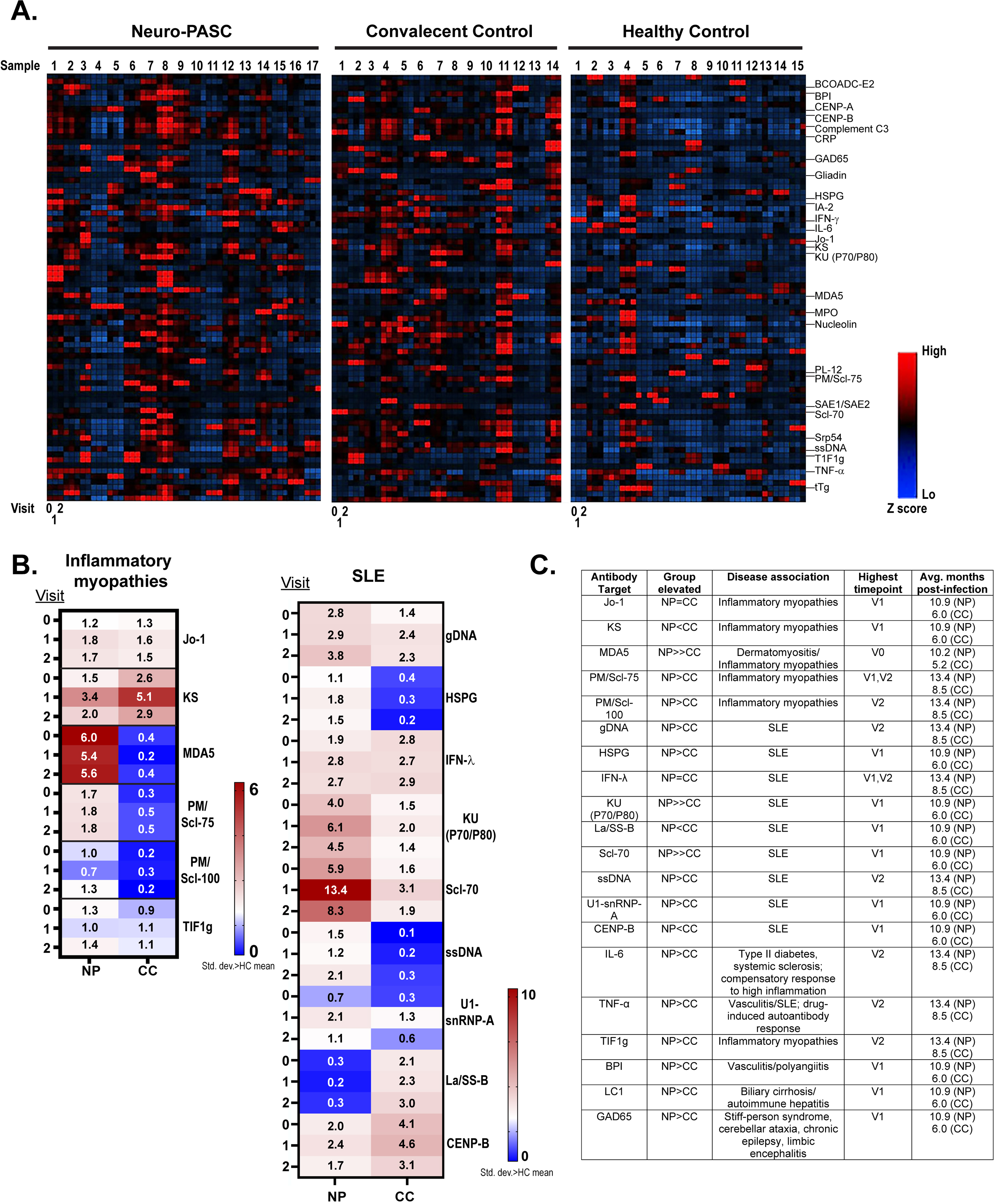
SARS-CoV-2 infection provokes sustained autoantibody elevation in Neuro-PASC patients and COVID convalescent controls. A.) Microarray of 80 IgG autoantibodies elevated in NP, CC, and HC groups across study visits up to 4 months post-mRNA booster dose. B.) Autoantibodies associated with autoimmune inflammatory myopathies (left) or SLE (right) remain elevated in NP and to a lesser extent CC subjects months after acute infection and vaccination. C.) Table summarizing data in B. Avg. months post-infection corresponds to timepoint (V0, 1, or 2) showing the greatest elevation in autoantibody titers.

### Prolonged elevations in antibodies linked to inflammatory myopathies and SLE are correlated with cognitive dysfunction in Neuro-PASC

Having identified that inflammatory myopathy and SLE-associated autoantibodies were enriched in Neuro-PASC patients, we determined whether titers were statistically different between groups. Though the frequency SLE-linked polyautoantibody responses did not differ between groups convalescent groups (Fig. S1), Neuro-PASC patients exhibited antibody responses several standard deviations above those of healthy controls at V2 with respect to the dermatomyositis antibody MDA5 and the SLE antibodies HSPG, KU (P70/P80), and ssDNA, unlike convalescent controls (Fig. 4A). Analysis of antibody titers similarly revealed significant elevations in anti-HSPG and anti-ssDNA titers only in Neuro-PASC patients at all times tested (Fig. 4B).

**Figure 4:**
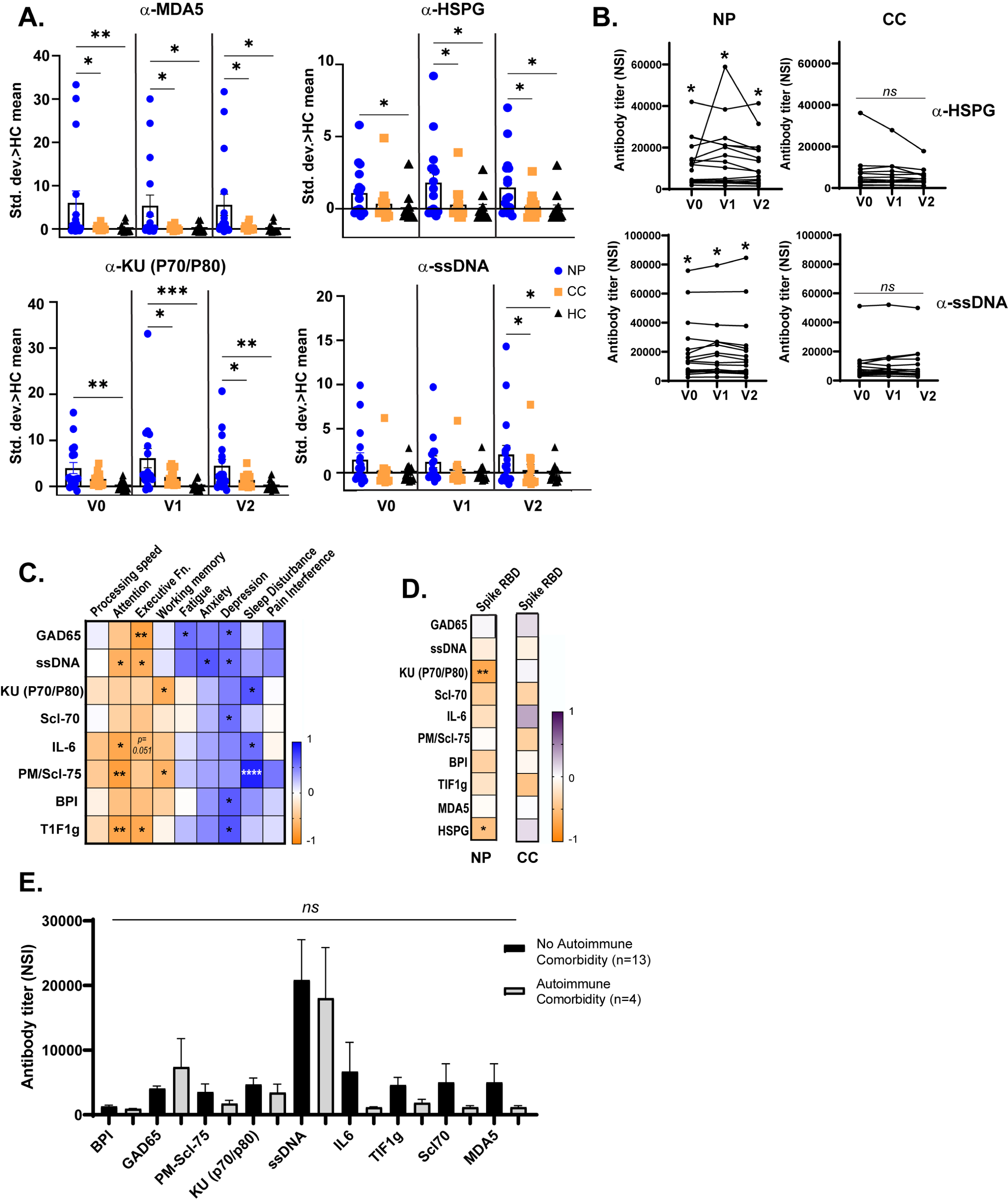
Autoantibodies associated with SLE and inflammatory myopathies are more elevated in Neuro-PASC patients and correlate with cognitive function. A.) NP patients have significantly higher antibody titers than CC against the dermatomyositis-associated antigen MDA5 and the SLE-associated antigens HSPG, KU (P70/P80), and ssDNA pre- and post-vaccination. B.) Longitudinal dynamics of HSPG and ssDNA autoantibodies in NP and CC subjects pre- and post-vaccination. C.) Higher titers of GAD65, ssDNA, and PM/Scl-75 and others correlate with lower scores in Attention, Executive Function, and Working Memory on NIH Toolbox cognitive tests and worse long COVID symptom severity in NP patients at V0 pre-boost. D.) Lower vaccine-elicited Spike RBD antibody responses are correlated with higher SLE-associated autoantibody titers in NP patients but not CC subjects at V2 post-vaccination. E.) SLE- and myositis-associated autoantibody titers are not affected by preexisting autoimmune comorbidities in NP patients. NSI: normalized signal intensity. *p<0.05; **p<0.01; ***p<0.001; ****p<0.0001 by one-way ANOVA with Fisher’s LSD posttest (A); one sample t test or Wilcoxon test where the mean/median was set to the average value of the HC group at each visit (B); or Spearman correlation (C-D).

Higher autoantibody responses in Neuro-PASC patients were also correlated with poor cognitive performance as determined by the NIH Toolbox cognition battery which measures cognitive domains known to be important for intellectual function, independence, and scholastic/workplace success^18^. High titers of the neuronal autoantibody against GAD65 strongly associated with poor results in Executive Function, while higher titers of the myositis antibody PM/Scl-75 correlated with low Attention scores (Fig. 4C). Higher autoantibody responses were similarly correlated with worse fatigue, anxiety, depression, and sleep disturbance in Neuro-PASC patients. Importantly, vaccine-elicited Spike RBD responses were decreased in the context of high HSPG and KU(P70/P80) titers only in Neuro-PASC patients but not convalescent controls at V2 (Fig. 4D). Preexisting diagnosis of autoimmune disease also did not affect autoantibody responses in Neuro-PASC patients (Fig. 4E). These results demonstrate that Neuro-PASC patients have higher autoantibody responses than COVID convalescent controls that correlate with cognitive dysfunction and symptom severity.

Our study is novel as it is the first to our knowledge to show that mild acute SARS-CoV-2 infection can lead to elevated autoantibody responses, with worse outcomes for patients with persistent neurologic symptoms of long COVID. We additionally show that mRNA vaccination does not significantly decrease autoantibody titers induced by SARS-CoV-2 infection, and that acute phase markers of inflammation do not have predictive value when evaluating autoantibody responses. Altogether, these data demonstrate that autoimmunity can be a consequence of mild COVID-19.

## Discussion

As up to 33% of people infected by SARS-CoV-2 experience persistent symptoms, it is estimated that over 30 million people in the US are affected by long COVID/PASC. While the precise mechanisms remain unclear, there is increasing evidence implicating humoral autoimmunity in the pathogenesis of Neuro-PASC after severe SARS-CoV-2 infection^10, 19^. However, there has been little focus on whether mild acute disease can impact autoimmunity, and how this differs upon vaccination in people with and without long COVID. We aimed to fill this knowledge gap by longitudinally examining how infection and vaccination affect autoantibody responses to 80 antigens in discrete populations of Neuro-PASC, COVID convalescent control, and healthy control subjects.

Over 94% of Neuro-PASC and 100% of convalescent controls in our study presented with mild acute disease. Despite this, 88% of Neuro-PASC and 85% of convalescent controls had statistically significant elevations in multiple autoantibodies after infection that did not decrease after vaccination. This was despite the fact that convalescent controls were mostly in their twenties with few pre-existing comorbidities. Our findings expand on recent studies showing that humoral autoimmunity is a contributing factor to neurologic symptoms of acute COVID-19 and PASC^20^. Importantly, studies to date have focused on the links between autoimmunity and acute disease severity because higher inflammation during severe disease can lead to bystander activation of autoreactive B and T cells or epitope spreading due to excessive tissue damage^21^. Indeed, the inability to resolve autoantibody elevation up to 1 year after severe acute infection has been linked to the persistence of fatigue, dyspnea, and other symptoms^9^.

However, there is historical and serological evidence that previous mild manifestations of coronavirus disease also led to persistent symptoms including cognitive impairment, muscle weakness, and dysautonomia during the Russian pandemic of 1889-1891^22, 23^. Additionally, 83% of COVID convalescents with mild or severe acute disease were found to have latent autoimmunity in the current pandemic^24^, demonstrating that elevated autoantibody titers may not always result in immediate symptoms^25^. It is possible that mild SARS-CoV-2 infection constitutes an environmental trigger that can induce latent autoimmunity^26^ in some people, which eventually manifests as symptomatic autoimmunity in Neuro-PASC patients. Our data also suggest that the development of latent autoimmunity in COVID convalescents may potentially put them at risk of PASC in case of recurrent SARS-CoV-2 infection. This is similar to studies showing that individuals were at a much higher risk for post-acute sequelae including neurologic and musculoskeletal manifestations after multiple infections compared with those who were only infected once^27^. We also identified such pronounced elevations of autoantibodies after mild acute disease in contrast to other reports^8, 28^ because our samples were collected at least 5 months and up to 1.5 years after acute infection as opposed to within 3 months of infection. This highlights the need for longitudinal studies to define autoimmune correlates and development of symptoms in Neuro-PASC patients and COVID convalescent controls.

Autoantibodies associated with SLE and inflammatory myopathies were more elevated in Neuro-PASC patients than convalescent controls and significantly correlated with lower cognitive performance and high symptom severity. PASC has been associated with anti-nuclear SLE antibodies after severe acute disease in other studies^29, 30^, but fewer studies have focused on antibodies related to inflammatory myopathies. Interestingly, high levels of the dermatomyositis antibody MDA5 were found in COVID-19 patients with severe acute disease who eventually succumbed to infection^31^. In our study, we detected significant elevations in MDA5 autoantibodies solely in Neuro-PASC patients and not COVID convalescent controls. This suggests that the MDA5 antibody may be a good candidate biomarker for PASC, but also raises questions about which pathogenic mechanisms Neuro-PASC and severe acute COVID-19 have in common. MDA5 is a cytosolic RIG-I-like receptor that senses viral RNA and ultimately lead to the production of type I interferons^32^. It is well-established that patients with moderate to severe acute COVID-19 have elevated autoantibodies against type I IFNs^33^, but our data suggest that autoantibodies against MDA5 upstream of type I IFN production may be involved in the pathogenesis of PASC. Though MDA5 antibody levels were significantly correlated with age, younger Neuro-PASC patients also had higher levels than COVID convalescent controls. Further studies are needed to determine whether SARS-CoV-2 can induce anti-MDA5 antibodies as an immune evasion strategy during persistent infection, which has also been linked to PASC^34–36^.

Previous studies have convincingly shown that SARS-CoV-2 mRNA vaccines do not *induce* autoantibody responses^37^, but they did not assess whether the vaccines can decrease existing humoral autoimmunity in COVID convalescents. We found that booster vaccine doses did not decrease autoantibody responses in Neuro-PASC patients or convalescent controls, suggesting that vaccination cannot be used to combat autoimmunity resulting from exposure to SARS-CoV-2. Most convalescent controls and a substantial portion of Neuro-PASC patients in our study also contracted breakthrough SARS-CoV-2 infections after receiving the primary series of mRNA vaccines. The observation that vaccinated patients developed autoantibody responses after infection demonstrates that vaccines may be unable to protect against autoimmune sequelae if a patient is infected with SARS-CoV-2. Furthermore, high levels of SLE autoantibodies were associated with lower vaccine-elicited Spike IgG response in Neuro-PASC patients, suggesting that this shift towards autoimmunity may lower the protective effect of vaccination in case of subsequent SARS-CoV-2 infection. These data highlight the need for additional strategies to decrease the spread of SARS-CoV-2 such as the development of vaccines that can prevent infection rather than decrease severity of acute disease alone. Longitudinal studies are also needed to determine whether autoimmune responses in COVID convalescents may lead to Neuro-PASC symptoms after recurrent SARS-CoV-2 infections despite vaccination. In conclusion, our work emphasizes the need to implement more public health strategies to decrease the spread of SARS-CoV-2 in the community.

### Lead contact statement

Further information and requests for resources and reagents should be directed to and will be fulfilled by the lead contact, Lavanya Visvabharathy.

### Materials Availability

This study did not generate new unique reagents.

### Data Availability

The full datasets generated in the current study are available from the corresponding author upon requests. The source data for all main figures are provided with this paper.

## Data Availability

The full datasets generated in the current study are available from the corresponding author upon requests.

## Acknowledgements

We thank the Team from Microarray and Immune Phenotyping Core at UT Southwestern Medical Center in Dallas, TX for their expert help with sample processing and autoantibody data generation. L.V. is supported by grant B48 from the philanthropic COVID research fund through Balvi Ops. P.P.M is supported by grants from the National Institute on Drug Abuse (NIDA, DP2DA051912) and from the National Institute of Biomedical Imaging and Bioengineering (NIBIB, U54EB027049). I.K. is supported by a grant from the National Institute of Aging (NIA, R01AG059291).

## Author contributions

Conceptualization L.V.; Investigation L.V., C.Z., Z.O., N.P., P.L., M.J.; Formal Analysis L.V. and C.Z.; Resources L.V., P.P.M., and I.K.; Data curation L.V.; Writing L.V. with feedback from all authors; Supervision L.V. and I.K.; Project Administration L.V.; Funding Acquisition: L.V. and I.K.

## STAR Methods

### Study participants, NIH Toolbox, and PROMIS-57 data collection

This study was approved by the Northwestern University Institutional Review Board, protocol number STU00212583. We enrolled consenting adult outpatients seen by neurologists in the Neuro-COVID-19 clinic at Northwestern Memorial Hospital from August2021-March 2022, including 17 Neuro-PASC patients with documented PCR^+^, rapid antigen^+^, or seropositive IgG results for SARS-CoV-2 prior to vaccination. All NP patients had lingering symptoms lasting >8 weeks. In parallel, we recruited 14 healthy COVID convalescents from the surrounding community who tested either PCR+ or seropositive for SARS-CoV-2 before vaccination but had no lingering symptoms lasting >4 weeks and 15 healthy controls who tested PCR- for SARS-CoV-2 and were also seronegative for IgG against SARS-CoV-2 Spike RBD prior to vaccination. All subjects were vaccinated with the primary series of either Pfizer or Moderna mRNA vaccines prior to the study. All study subjects remained living throughout the period of observation. Heparinized blood samples were collected one time from each subject at an average of 21-40 weeks post-symptom onset (as in Fig. 1B). Other demographic information, including preexisting comorbidities, is contained in Fig. 1B. NP patients completed a cognitive function evaluation in the clinic coincident or near the date of their blood sample acquisition with the National Institutes of Health (NIH) Toolbox v2.1 instrument, including assessments of processing speed (pattern comparison processing speed test); attention and executive memory (inhibitory control and attention test); executive function (dimensional change card sort test); and working memory (list sorting working memory test)^18^. PROMIS-57 patient-reported quality of life assessments were administered to Neuro-PASC patients at the time of clinic visit. Both PROMIS-57 and NIH Toolbox results are expressed as T-scores with a score of 50 representing the normative mean/median of the US reference population and a standard deviation of 10. Toolbox results are adjusted for age, education, gender, and race/ethnicity. Lower cognition T-scores indicate worse performance while higher fatigue, depression, anxiety, or pain interference T-scores indicate greater symptom severity.

### Plasma collection

30mL of venous blood from study volunteers was collected in blood collection tubes containing sodium heparin from BD Biosciences. Whole blood was layered on top of 15mL of Histopaque 1077 (Sigma-Aldrich) in 50mL Leucosep blood separation tubes (Greiner Bio-One) and spun at 1000g for 18min at RT. Plasma was collected and stored at -80°C until further use.

### IgG Spike RBD/CRP ELISA

Spike RBD-specific total antibody titers were measured by ELISA as described previously^38^. In brief, 96-well flat-bottom MaxiSorp plates (Thermo Scientific) were coated with 1 µg/ml of Spike RBD for 48 hr at 4°C. Plates were washed three times with wash buffer (PBS + 0.05% Tween 20). Blocking was performed with blocking solution (PBS + 0.05% Tween 20 + 2% bovine serum albumin), for 4 hr at room temperature. 6 µl of sera was added to 144 µl of blocking solution in the first column of the plate, 1:3 serial dilutions were performed until row 12 for each sample, and plates were incubated for 60 min at room temperature. Plates were washed three times with wash buffer followed by addition of secondary antibody conjugated to horseradish peroxidase, goat anti-human IgG (H + L) (Jackson ImmunoResearch) diluted in blocking solution (1:1000) and 100 µl/well was added and incubated for 60 min at room temperature. After washing plates three times with wash buffer, 100 µl/well of Sure Blue substrate (SeraCare) was added for 1 min. Reaction was stopped using 100 µl/well of KPL TMB Stop Solution (SeraCare). Absorbance was measured at 450 nm using a Spectramax Plus 384 (Molecular Devices). SARS-CoV-2 RBD protein used for ELISA was produced at the Northwestern Recombinant Protein Production Core by Dr. Sergii Pshenychnyi with plasmids produced under HHSN272201400008C and obtained from BEI Resources, NIAID, NIH: Vector pCAGGS containing the NR-52394 Spike receptor binding domain (RBD) protein region. Plasma CRP levels were quantified using the CRP Human ELISA kit (ThermoFisher).

### Autoantibody profiling

Plasma IgG reactivities against 80 discrete autoantigens were assessed using a microfluidic antigen array available through the Microarray and Immune Phenotyping Core at the UT Southwestern Medical Center. In brief, 10uL of plasma was spotted in triplicate on a 16-pad FAST slide. Autoantibodies binding to arrayed antigens were detected with Cy3-labeled anti-human IgG. Arrays were scanned with the GenePix 4400A Microarray Scanner (Molecular Devices) and images in the array were converted to Genepix Report file (GPR) with Genepix Pro7.0 software (Molecular Device). The averaged fluorescent signal intensity of each antigen was subtracted by local background and the PBS control signal and normalized to internal controls to obtain the normalized fluorescence intensity (NSI) value.

### Quantification and Statistical Analysis

Statistical tests to determine significance are described in figure legends and conducted largely in Prism (GraphPad). Z scores for autoantibody heatmaps were calculated in R. Clinical data were collected and managed using REDCap electronic data capture tools hosted at Northwestern University Feinberg School of Medicine. All error bars on figures represent values ± SEM.

### Study approval

This study was approved by the Northwestern University Institutional Review Board (Koralnik Lab, IRB STU00212583). Informed consent was obtained from all enrolled participants. Samples were de-identified before banking.

## Supplemental Figure Titles & Legends

**Figure S1: Total numbers of significantly elevated autoantibodies do not differ between Neuro-PASC and convalescent control groups.**

A, C). Titers of antibodies linked to SLE- and inflammatory myopathies were transformed into standard deviations above the mean for unexposed healthy controls at each visit. Data represent total number of antibodies elevated at least >2SD compared with mean for healthy controls for each patient. B, D). Autoantibody specificities included in analysis for A, C.

**Figure S2: Elevated anti-cytokine, liver antigen, and neuronal antigen-associated autoantibodies in NP patients.**

Autoantibodies associated with neurological dysfunction, vasculitis, biliary cirrhosis, and excessive inflammation are more highly elevated in NP patients. Heatmap calculated based on standard deviations above the mean for HC subjects at each visit.

**Figure S3: Autoantibody correlations with demographic data**

A.) Autoantibody titers do not correlate with age in NP and CC subjects (left panel) except for MDA5 (right panel). B.) Autoantibody titers do not correlate with time since infection at V0 pre-boost (top panel) except for MDA5 levels which increase over time in both NP and CC subjects (bottom panel). C.) Autoantibody responses are not impacted by sex or pre-existing cardiac or metabolic comorbidities, but some SLE-associated species are positively correlated with preexisting autoimmune disease. *p<0.05; **p<0.01 by simple linear regression (A,B bottom panels), or multiple linear regression (A,B top panels; E).

**Figure S4: CRP levels cannot predict elevated autoantibody titers in subjects who have been infected with SARS-CoV-2.**

Polyautoimmunity pre- and post-vaccination are not linked to levels of the acute-phase inflammatory marker CRP, age, or the presence of comorbidities.

