## Supplemental material for "Autoantibody production is enhanced after mild SARS-CoV-2 infection despite vaccination in individuals with and without long COVID"

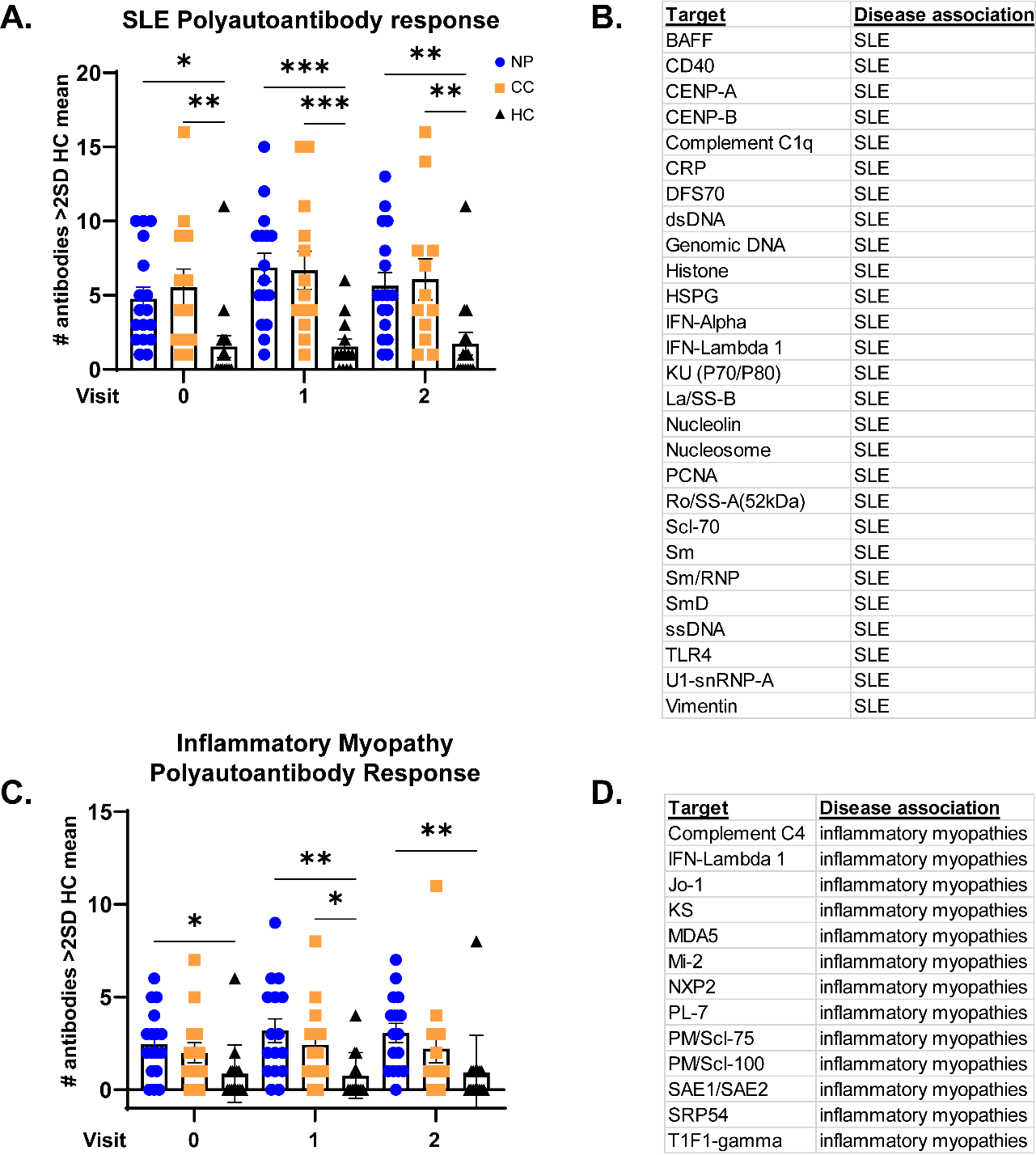

**Figure S1: Total numbers of significantly elevated autoantibodies do not differ between Neuro-PASC and convalescent control groups.**

A, C). Titers of SLE- and inflammatory myopathy-associated antibodies were transformed into standard deviations above the mean for unexposed healthy controls at each visit. Data represents total number of antibodies elevated at least >2SD compared with healthy controls for each patient. B, D). Autoantibody specificities included in analysis for A, C.

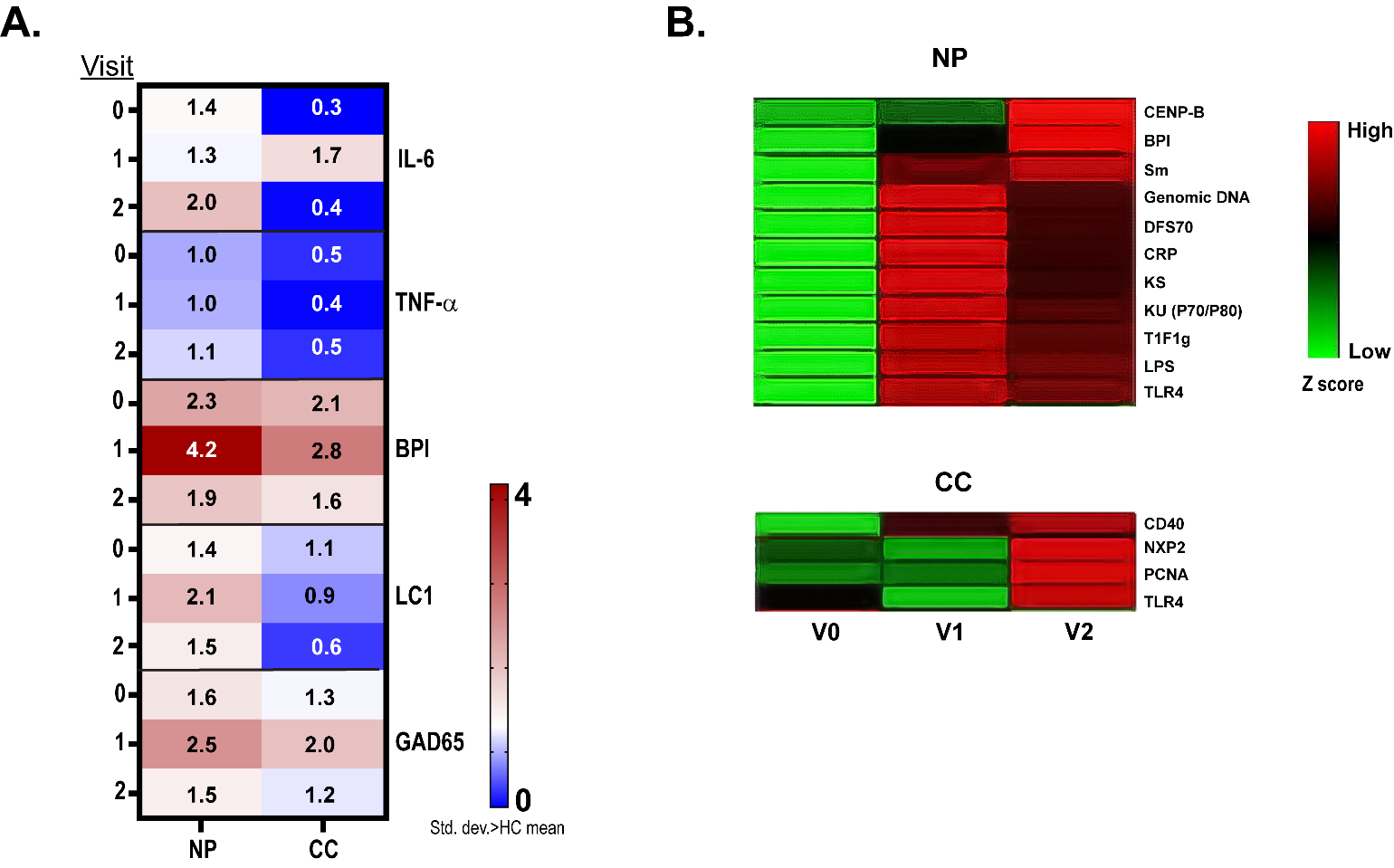

**Figure S2: Elevated anti-cytokine, liver antigen, and neuronal antigen-associated autoantibodies in NP patients.**

Autoantibodies associated with neurological dysfunction, vasculitis, biliary cirrhosis, and excessive inflammation are more highly elevated in NP patients. Heatmap calculated on the basis of standard deviations above the mean for HC subjects at each visit.

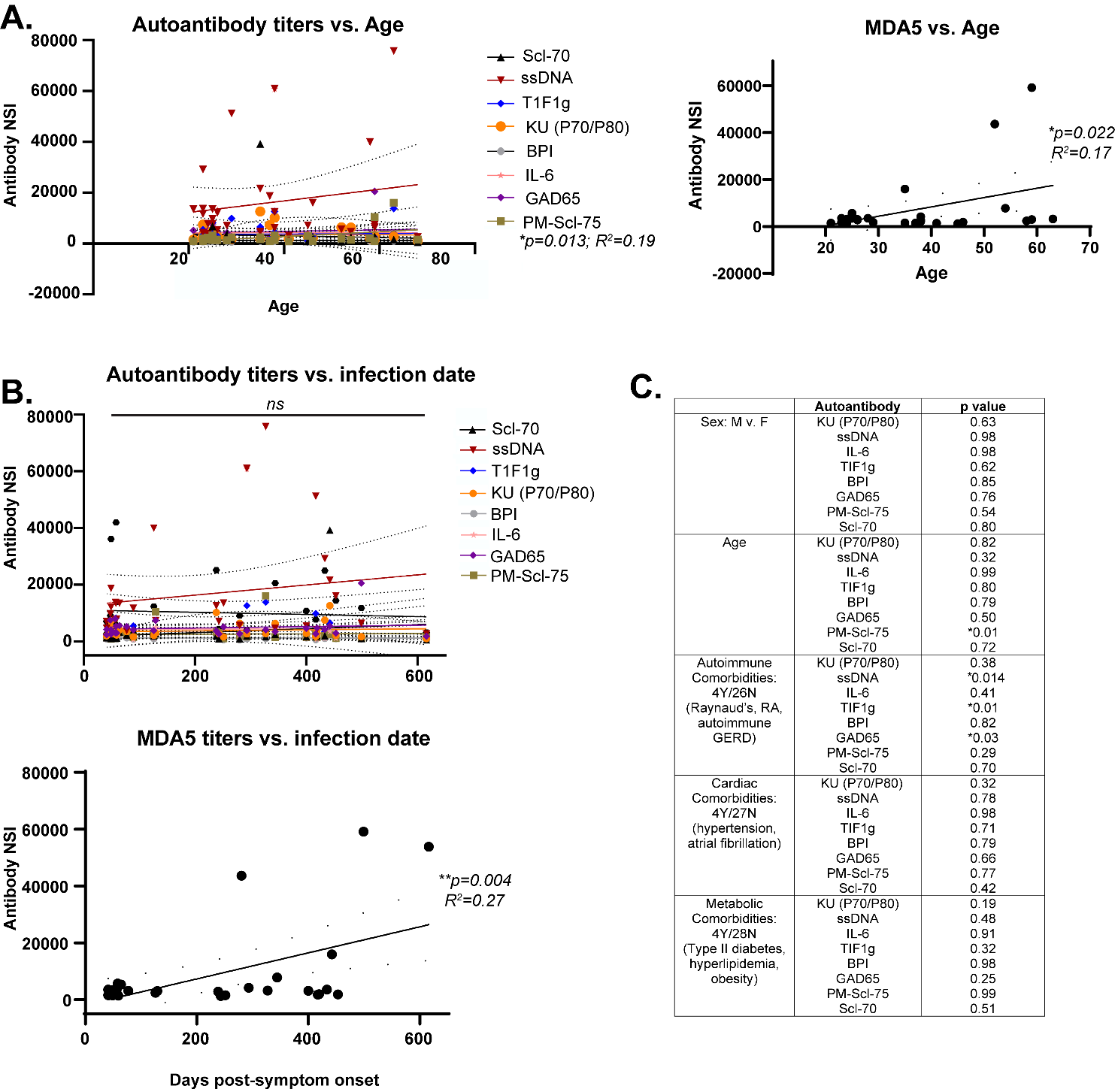

**Figure S3: Autoantibody correlations with demographic data**

A.) Autoantibody titers do not correlate with age in NP and CC subjects (right panel) except for MDA5 (left panel). B.) Autoantibody titers do not correlate with time since infection at V0 pre-boost (top panel) except for MDA5 levels which go up over time in both NP and CC subjects (bottom panel). C.) Autoantibody responses are not impacted by sex or pre-existing cardiac or metabolic comorbidities, but some SLE-associated species are positively correlated with preexisting autoimmune disease. *p<0.05; **p<0.01 by simple linear regression (A,B bottom panels), or multiple linear regression (A,B top panels; E).

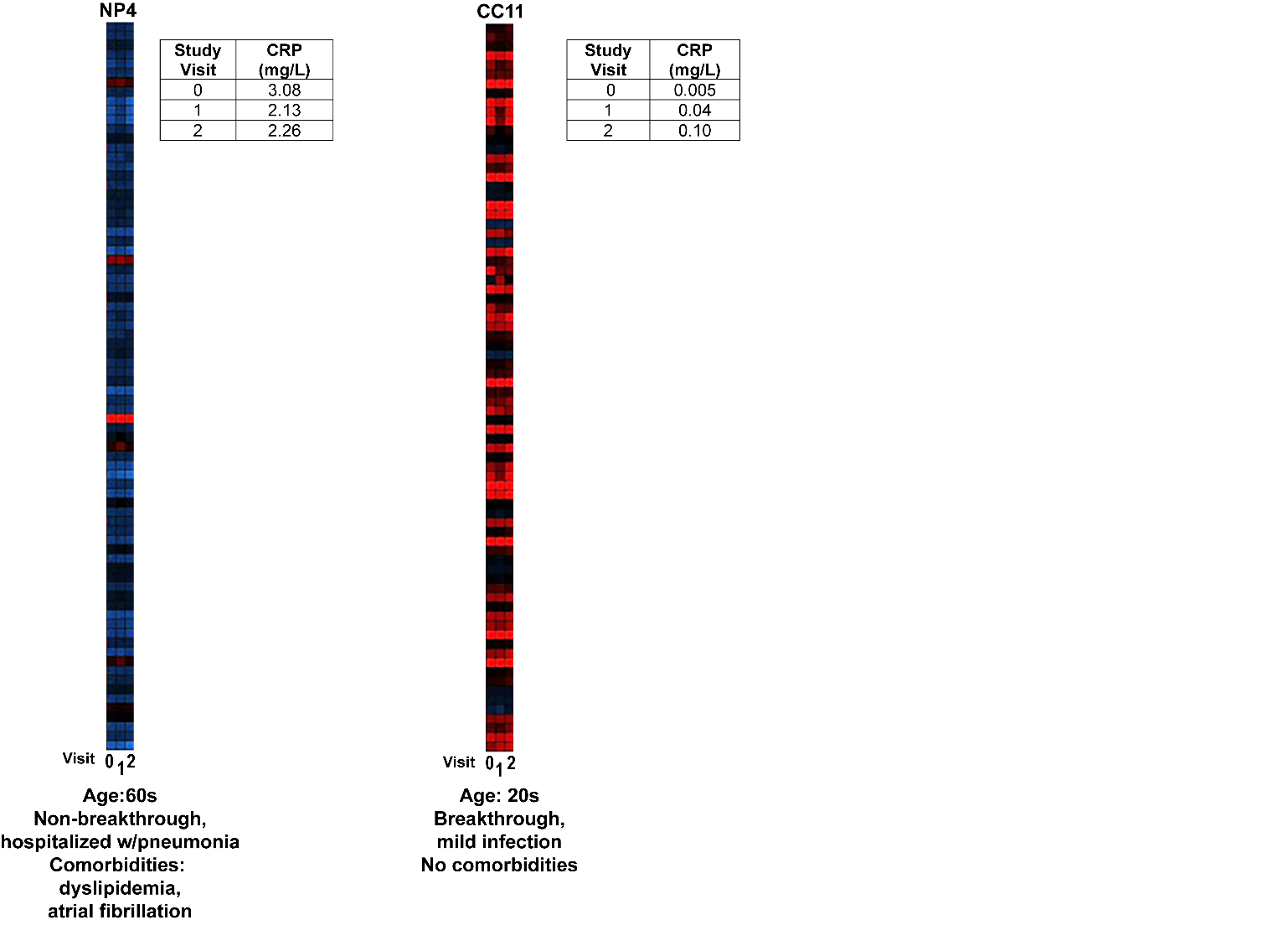

**Figure S4: CRP levels cannot predict elevated autoantibody titers in subjects who have been infected with SARS-CoV-2.**

Autoantibody titers pre- and post-vaccination are not linked to levels of the acute-phase inflammatory marker CRP or presence of rheumatoid factor autoantibodies.

**Table S1: Autoantibodies screened in microarray**

| **Protein** | **Autoantibody symbol** |
| --- | --- |
| Aggrecan | Aggrecan |
| Alanyl-tRNA Synthetase (PL-12) | PL-12 |
| Asparaginyl-tRNA Synthetase (KS) | KS |
| Bactericidal/permeability-increasing protein (BPI) | BPI |
| ß2-Glycoprotein 1 | ß2-Glycoprotein 1 |
| BCOADC-E2 | BCOADC-E2 |
| Cardiolipin | Cardiolipin |
| Centromere Protein A (CENP-A) | CENP-A |
| Centromere Protein B (CENP-B) | CENP-B |
| Chondroitin sulfate | Chondroitin sulfate |
| Collagen III | Collagen III |
| Collagen V | Collagen V |
| Complement C3 | Complement C3 |
| Complement C4 | Complement C4 |
| Complement component C1q receptor (C1q) | Complement C1q |
| Cytochrome c | Cytochrome c |
| Cytochrome P450 2D6 (LKM 1; ng) | LKM 1 |
| DFS70 | DFS70 |
| DNA Topoisomerase I (Scl-70) | Scl-70 |
| Double-stranded DNA (dsDNA) | dsDNA |
| Elastin | Elastin |
| Fibrinogen Type I-S | Fibrinogen Type I-S |
| Formiminotransferase Cyclodeaminase (LC1) | LC1 |
| Genomic DNA | Genomic DNA |
| Gliadin | Gliadin |
| Glomerular Basement Membrane (GBM) | GBM |
| Glutamate Decarboxylase 65 kDa (GAD65; ng) | GAD65 |
| GP2 | GP2 |
| GP210 | GP210 |
| Heparan sulfate proteoglycan(HSPG) | HSPG |
| Histidyl-tRNA Synthetase (Jo-1) | Jo-1 |
| Histone | Histone |
| Human CD 40 | Human CD 40 |
| IA-2 (ICA 512)Insulinoma-associated protein (IA-2) | IA-2 |
| Insulin | Insulin |
| Intrinsic Factor (IF) | IF |
| KU (P70/P80) | KU (P70/P80) |
| La/SS-B | La/SS-B |
| Laminin | Laminin |
| M2 | M2 |
| MDA5 | MDA5 |
| Mi-2 | Mi-2 |
| Myelin basic protein (MBP) | MBP |
| Myeloperoxidase (MPO) | MPO |
| Myosin | Myosin |
| Nucleolin | Nucleolin |
| Nucleosome | Nucleosome |
| Nup 62 | Nup 62 |
| NXP2 also known as MORC3 | NXP2 |
| OGDC-E2 | OGDC-E2 |
| PDC-E2 | PDC-E2 |
| PM/Scl 100 | PM/Scl 100 |
| PM/Scl-75 | PM/Scl-75 |
| Proliferating Cell Nuclear Antigen (PCNA) | PCNA |
| Proteinase 3 (PR3) | PR3 |
| Proteoglycan | Proteoglycan |
| Ribosomal Phosphoprotein P0 | P0 |
| Ribosomal Phosphoprotein P2 | P2 |
| Ro/SS-A (52 kDa) | Ro/SS-A (52 kDa) |
| Ro/SS-A (60 Kda) | Ro/SS-A (60 Kda) |
| SAE1/SAE2 | SAE1/SAE2 |
| Sm | Sm |
| Sm/RNP | Sm/RNP |
| SmD | SmD |
| SmD1 | SmD1 |
| SP100 | SP100 |
| Fibrinogen IV | Fibrinogen IV |
| SRP54 | SRP54 |
| ssDNA | ssDNA |
| T1F1 gamma | T1F1 gamma |
| Threonyl-tRNA Synthetase (PL-7) | PL-7 |
| Thyroglobulin | Thyroglobulin |
| Thyroid Peroxidase (TPO) | TPO |
| Tissue Transglutaminase (tTG) | tTG |
| TNF-alpha | TNF-alpha |
| U1-snRNP 68/70 kDa | U1-snRNP 68/70 kDa |
| U1-snRNP A | U1-snRNP A |
| U-snRNP B/B' | U-snRNP B/B' |
| Vimentin | Vimentin |
| Vitronectin | Vitronectin |

**Table S2: Patient demographics & vaccination information**

| **Sample** | **Age** | **Sex** | **Race** | **Vaccine doses** | **Vaccine brand** | **Breakthrough?** |
| --- | --- | --- | --- | --- | --- | --- |
| NP1 | 20s | F | Asian | 3 | Pfizer 3x | Y |
| NP2 | 50s | F | Black | 3 | Pfizer 3x | N |
| NP3 | 30s | F | Hispanic | 3 | Pfizer 3x | N |
| NP4 | 50s | F | Asian | 3 | Pfizer 3x | N |
| NP5 | 60s | M | White | 3 | Pfizer 3x | N |
| NP6 | 50s | F | Asian | 3 | Pfizer 3x | N |
| NP7 | 60s | M | White | 3 | Pfizer 3x | N |
| NP8 | 30s | M | White | 3 | Pfizer 3x | N |
| NP9 | 30s | F | Black | 3 | Pfizer 3x | N |
| NP10 | 50s | F | Hispanic | 3 | Pfizer 3x | Y |
| NP11 | 20s | F | Asian | 3 | Moderna 3x | Y |
| NP12 | 40s | F | White | 3 | Moderna 3x | N |
| NP13 | 50s | M | White | 3 | Pfizer 3x | Y |
| NP14 | 30s | F | Asian | 2 | Janssen/Moderna | Y |
| NP15 | 40s | F | White | 3 | Pfizer 3x | N |
| NP16 | 20s | M | Other | 3 | Pfizer 3x | Y |
| NP 17 | 20s | F | Hispanic | 3 | Pfizer 3x | N |
| CC1 | 20s | F | Asian | 3 | Pfizer 3x | Y |
| CC2 | 20s | F | White | 3 | Pfizer 3x | N |
| CC3 | 20s | M | White | 3 | Pfizer 3x | Y |
| CC4 | 20s | M | Asian | 3 | Moderna 3x | Y |
| CC5 | 20s | M | White | 3 | Pfizer 3x | Y |
| CC6 | 20s | M | White | 3 | Moderna 3x | Y |
| CC7 | 20s | M | White | 3 | Pfizer 3x | Y |
| CC8 | 20s | M | Black | 3 | Pfizer 3x | N |
| CC9 | 20s | M | Hispanic | 3 | Pfizer 3x | Y |
| CC10 | 30s | M | Asian | 3 | Moderna 3x | Y |
| CC11 | 20s | F | Asian | 3 | Pfizer 3x | Y |
| CC12 | 30s | F | Hispanic | 3 | Pfizer 3x | Y |
| CC13 | 40s | M | Hispanic | 3 | Moderna 3x | Y |
| CC14 | 20s | F | Black/Asian | 3 | Pfizer 3x | Y |
| HC1 | 50s | M | White | 3 | Pfizer 3x | NA |
| HC2 | 20s | F | Asian | 3 | Pfizer 3x | NA |
| HC3 | 20s | M | White | 3 | Pfizer 3x | NA |
| HC4 | 20s | M | Asian | 3 | Pfizer 3x | NA |
| HC5 | 20s | F | White | 3 | Pfizer 3x | NA |
| HC6 | 20s | M | Asian | 3 | Pfizer 3x | NA |
| HC7 | 40s | M | White | 3 | Moderna 3x | NA |
| HC8 | 60s | F | White | 3 | Pfizer 3x | NA |
| HC9 | 20s | F | Asian | 3 | Pfizer 3x | NA |
| HC10 | 60s | F | Other | 3 | Pfizer 3x | NA |
| HC11 | 40s | F | White | 3 | Pfizer 3x | NA |
| HC12 | 30s | M | Asian | 3 | Moderna 3x | NA |
| HC13 | 30s | F | Asian | 3 | Moderna 3x | NA |
| HC14 | 20s | F | Asian | 3 | Pfizer 3x | NA |
| HC15 | 30s | F | Other | 3 | Pfizer 3x | NA |
